# A Simplified Bayesian Analysis Method for Vaccine Efficacy

**DOI:** 10.1101/2020.12.07.20244954

**Authors:** Carlo Graziani

## Abstract

We describe a simplified Bayesian analysis of vaccine trial data, in which a reparametrization of the Poisson likelihood leads to a factorization in which the protective vaccine efficacy *VE*_*S*_ and the nuisance parameter appear in different factors. As a consequence the posterior density acquires a factorized form, and marginalization over the nuisance parameter is trivial. Estimates of *VE*_*S*_ accordingly become a matter of simple manipulations of one-dimensional posterior probability densities. We demonstrate the method using the publically-released data on the efficacy of three vaccines agains SARS-CoV-2: the final Phase III data from the Pfizer/BioNTech and Moderna mRNA vaccines and the interim data released for the Sputnik V adenovirus-based vaccine.

## 1. Introduction

A key parameter of a vaccine against an infectious disease is the *protective vaccine efficacy*, usually denoted *VE*_*S*_. This parameter is intended to represent the reduction due to the vaccine of the chance of contracting the disease, and is typically estimated using controlled double-blind clinical trials [1]. Simple point-estimates of *VE*_*S*_ may be produced by person-time exposure-weighted ratios of vaccine group incidence to placebo group incidence. Such estimates are known to be affected by correctable biases [2]. From a statistical perspective, however, interval estimates are more valuable than point estimates, as they yield information concerning the uncertainty in the estimates. Frequentist methods for estimating confidence regions have been proposed [e.g. 3, 4] and compared for relative conservativeness and accuracy of their coverage [5, 6].

Bayesian credible region estimation methods have also been proposed [7, 8]. Such methods are straightforward to implement and interpret, although they usually raise issues of choice of prior distribution over the parameters — both over *VE*_*S*_ and over another “nuisance” parameter, often the disease incidence in the placebo group. In [7] the prior density is chosen to be a product of log-normals, with “uninformative” widths. The work described in [8] examines the use of reference priors, as a means of exhibiting principled notion of “uninformativeness” [9]. It is worth noting that one reason for concern with prior choice is the extra degree of freedom represented by the nuisance parameter, since the posterior density must be marginalized over the nuisance parameter in order to obtain inferences that concern *VE*_*S*_ alone. It is a matter of due-diligence to demonstrate that such inferences are robust despite different prior choices, that is, that credible regions and point estimates suffer negligible variation across plausible families of prior densities. In many cases this desirable property of robustness is expected *a priori* for parameter estimation in the limit of abundant data, since in this case the likelihood is typically so sharply peaked that all priors of interest are for all intents and purposes constant in the parameter-space region where the mass of the posterior is concentrated [10]. However, the higher the dimension of the parameter space in which parameter inference tasks are performed, the greater the risk of loss of robustness to prior choice. In particular, if the nuisance parameter could be summarily ejected from the Bayesian estimation of *VE*_*S*_, the resulting estimates should be expected to be more robust against prior choice, as well as simpler to obtain.

In this article we briefly describe a reparametrization of the Poisson likelihood that permits precisely this simplification: the choice of a new, reparametrized nuisance parameter *λ* — the total expected infections across both vaccine and placebo group — results in a *factorization* of the likelihood into a factor depending only on *VE*_*S*_ and a second factor depending only on *λ*. As a result, one obtains a simple, intuitive result for the *reduced likelihood ℒ*_*e*_ (*VE*_*S*_), which in turn permits very simple inferences about credible regions, that are better insulated against variations due to prior choice.

We describe the method in §2. In §3 we exhibit it’s use in a study that compares the efficacy of three recently-announced vaccines agains SARS-CoV-2: The Pfizer/BioNTech mRNA vaccine [11], the Moderna mRNA vaccine [12], and the Sputnik V adenovirus-based vaccine [13]. We supply some final discussion in §4.

## 2. The Factorized Likelihood

We assume that infections in the placebo and vaccine samples are well-described by Poisson models with different means *µ*_*P*_ and *µ*_*V*_, respectively. For the sake of notational compactness, we will denote the protective vaccine efficacy *VE*_*S*_ by the symbol *e*. We introduce a model for *e* by assuming that

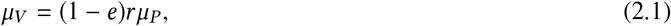

where *r* is the vaccine-to-placebo ratio of person-times at risk – that is, if the vaccine and placebo groups represent *P*_*V*_ and *P*_*P*_ person-times of exposure to the pathogen respectively, then *r* ≡ *P*_*V*_ / *P*_*P*_. If the infection numbers recorded during the trial are *N*_*V*_ and *N*_*P*_ respectively for the vaccine and placebo groups, then the likelihood under the model is the product of the Poisson likelihoods for the two groups, that is

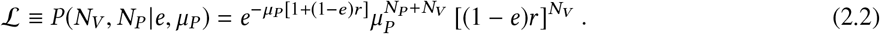

This form of the likelihood entangles the nuisance parameter *µ*_*P*_ with the quantity of interest, the efficacy *e*, so that after introducing a prior distribution and using Bayes’ theorem, the posterior distribution over *e, µ*_*P*_ is similarly entangled. Rather than submitting to the necessity of explicitly marginalizing the posterior over *µ*_*P*_, we may introduce a factorized form of the likelihood by means of a change of variables. We introduce the parameter *λ*, defined as

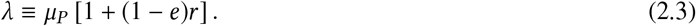

The new parameter *λ* is the expected number of overall infections, summed over the two groups — this is clear, since the sum of two Poisson random variables is itself a Poisson random variable whose mean is the sum of their respective means. We may use Equation (2.3) to substitute *λ* for *µ*_*P*_ in Equation (2.3). The resulting likelihood is

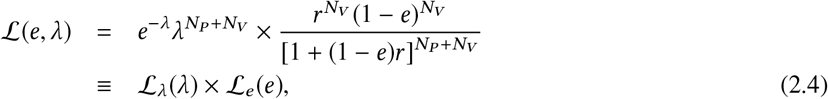

where we have introduced the *reduced likelihood* for *e*,

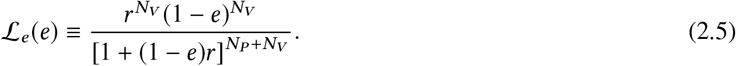

The advantage of the form of the likelihood in Equation (2.3) is that the dependence on *e* is factorized from the dependence on *λ*. If we now introduce a factorized prior *π*_*λ*_ (*λ*) ×*π*_*e*_ (*e*), then we obtain a factorized posterior distribution. Marginalization over *λ* is trivial, and we obtain for the marginalized posterior the formula

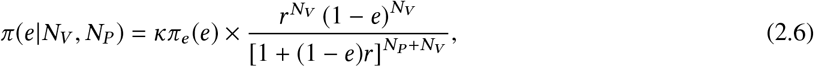

where *κ* is a normalization constant.

From this point of the discussion, we assume, for simplicity, a proper uniform prior *π*_*e*_ (*e*) = 1 on the domain *e* ∈ [0, 1]. In §3.3 we will re-examine the effect of prior choice on this model.

One may easily show that if *N*_*V*_ 0 then the posterior in Equation (2.3) has a maximum at 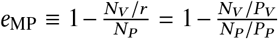, which accords with intuition – the relative incidence 1 − *e*_MP_ is given by the ratio of the person-time adjusted infection rates. On the other hand, when *N*_*V*_ = 0 the posterior increases monotonically, attaining a domain maximum at *e*_MP_ = 1. This also accords with intuition: if no vaccinated subjects become infected, the most probable efficacy is 1.0 — perfect efficacy. Of course, in this case the distribution may still have substantial mass at much lower efficacies, depending on the value of *N*_*P*_.

The normalization constant *κ* may be calculated by integrating Equation (2.3) with respect to *e*, in the interval [0, 1]. This may be done by the change of variables *e* →*g* = 1 + (1 − *e*) *r*, expanding the resulting integrand using the binomial theorem. The result is

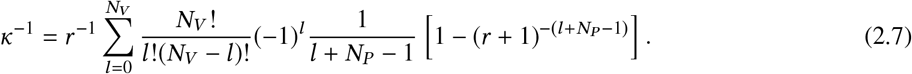

The usage of the formula for the posterior in Equation (2.3) is very straightforward. One may simply bin the interval 0, 1 into *N*_Samp_ equally-spaced sample points, compute the factorized likelihood at each point (multiplying by a non-uniform prior if so desired), and compute the normalization *κ* either by Equation (2.3) (for a uniform prior) or by a straightforward numerical quadrature. Then, after normalization, various desired interval estimates (such as credible regions, or upper/lower bounds of prescribed probability) can be obtained by sorting the array containing the 1-dimensional discretized posterior over *e*, and processing it as appropriate.

We now demonstrate the use of these procedures using public data from SARS-Cov-2 vaccine trials.

## 3. Bayesian Analysis of SARS-CoV-2 Vaccine Trial Data

At the time of this writing (early December 2020), the final data from the Phase III trials of the mRNA vaccines against SARS-CoV-2 from Pfizer/BioNTech [11] and from Moderna [12] are available from public information, although not yet from scientific journal articles. The two data releases include results for entire trials, and for subsamples of infections leading to “severe” COVID-19 symptoms. In addition, preliminary data from the Sputnik V vaccine trials are also available [13]. The data are summarized in Table 1.

**Table 1:** SARS-CoV-2 Vaccine Trial Data

| Trial | Vaccine Group Infections | Placebo Group Infections | $r$ |
| --- | --- | --- | --- |
| Pfizer/BioNTech (Overall) | 8 | 262 | 1.0 |
| Pfizer/BioNTech (Severe) | 1 | 9 | 1.0 |
| Moderna (Overall) | 11 | 185 | 1.0 |
| Moderna (Severe) | 0 | 30 | 1.0 |
| Sputnik V (Preliminary) | 8 | 31 | 3.0 |

The results of applying the reduced likelihood Bayesian analysis (with a uniform prior) to these samples and sub-samples are summarized in Table 2, and displayed in the panels of Figure 3.1. The figures display the posteriors over the efficacy *e* (blue solid line), the maximum-posterior efficacies (cyan dashed line), equal-posterior-density 90% credible regions (red solid region), and 99% lower bounds (hatched regions). These are all obtained by straightforward application of the method described in §2, implemented in a short *Python* script that makes use only of the *Numpy* [14] and *Matplotlib* [15] modules. The code is available for public download under an MIT license at https://github.com/CarloGraziani/BayesVaccineEfficacy.

**Figure 3.1:**
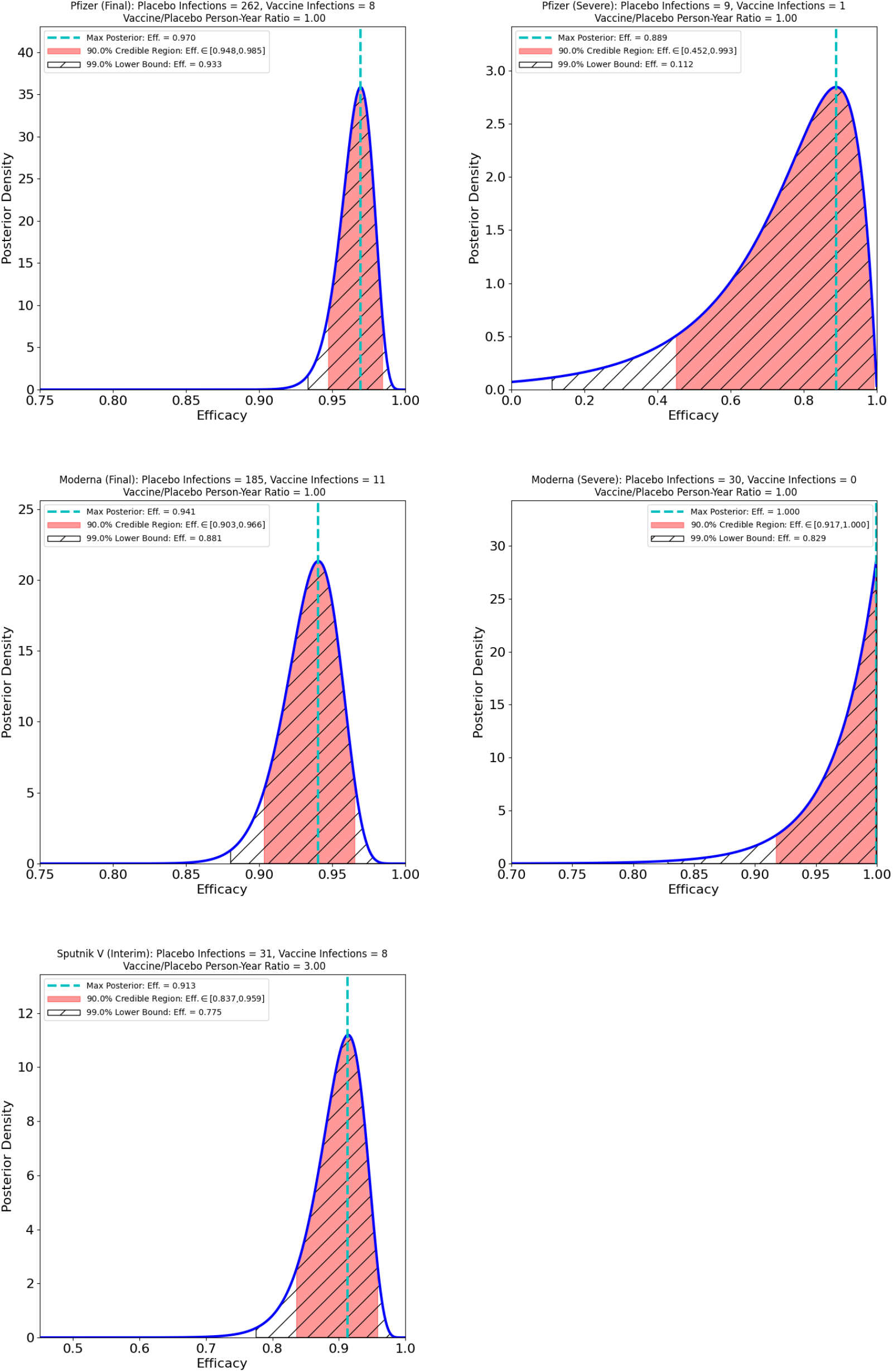
Efficacy estimates: Maximum a posteriori estimates (dashed line), 90% credible regions (solid shaded regions), and 99% lower bounds (hatched regions). Top: Pfizer/BioNTech, overall (left) and severe (right). Middle: Moderna overall (left) and severe (right). Bottom: Interim Sputnik V data.

**Table 2:** Summary of Bayesian efficacy estimation for SARS-CoV-2 vaccine trials.

| Trial | Max. Posterior | 90% Credible Region | 99% Lower Bound |
| --- | --- | --- | --- |
| Pfizer/BioNTech (Overall) | 0.970 | [0.948, 0.985] | 0.933 |
| Pfizer/BioNTech (Severe) | 0.889 | [0.452, 0.993] | 0.112 |
| Moderna (Overall) | 0.941 | [0.903, 0.966] | 0.881 |
| Moderna (Severe) | 1.0 | [0.917, 1.0] | 0.829 |
| Sputnik V (Preliminary) | 0.913 | [0.837, 0.959] | 0.775 |

We now discuss these results in more detail.

### 3.1. Overall Efficacy

From the left-hand side panels of Figure 3.1, we may note first that, as expected, the preliminary data released for the Sputnik V vaccine is much less constraining of the efficacy than is the final Phase III data for the Pfizer/BioNTech and for the Moderna vaccines. The reason is obvious, but instructive: the likelihood, and hence the posterior, gathers concentration with increasing *N*_*P*_, *N*_*V*_, and these numbers are, at this time, lower for the Sputnik V vaccine trials than for the other two trials.

A comparison of the Pfizer/BioNTech vaccine and the Moderna vaccine show broadly consistent results. Pfizer/BioNTech shows more posterior concentration (90% credible region: *e* ∈ [0.948, 0.985], 99% lower bound: *e* = 0.933), than does Moderna (90% credible region: *e* ∈ [0.903, 0.966 99] % lower bound: *e* = 0.881), but the intervals overlap enough to suggest that it is entirely possible on present data that the efficacies of the two vaccines are the same.^1^

These results are also consistent with the announced efficacy estimates from the (as of this date unpublished) statistical analyses carried out by Pfizer/BioNTech (estimated efficacy: 95% [11]) and by Moderna (estimated efficacy: 94.1% [12]). The Bayesian estimate for the efficacy of the Sputnik V vaccine is also consistent with the announced estimate from preliminary data (91.4% [13]), although as pointed out above this data is less constraining of the final result than is the data for the other two trials.

### 3.2. “Severe” Sub-Samples

An analysis of the “severe” case data suggests that the recruitment programs for the Pfizer/BioNTech and the Moderna trials were significantly different, and that the Moderna trial recruited populations at higher risk of severe COVID-19 symptoms. According to the published data [12] the Moderna trial featured 30,000 subjects, divided equally into placebo control and vaccine groups. Of the 15,000 control subjects, 30 developed “severe” symptoms. On the other hand, the Pfizer/BioNTech trial [11] featured 43,661 participants, half of whom were in the control group, and only 9 of these developed “severe” symptoms. On this data, it seems clear that the populations in the two trials must have been selected quite differently, since the control group incidence of severe COVID-19 appears to have been about 4.5 times higher for the Moderna trial than for the Pfizer/BioNTech trial. This is possibly explained by the released information about the Moderna trial [12], which asserts “The COVE study includes more than 7,000 Americans over the age of 65. It also includes more than 5,000 Americans who are under the age of 65 but have high-risk chronic diseases that put them at increased risk of severe COVID-19, such as diabetes, severe obesity and cardiac disease. These medically high-risk groups represent 42% of the total participants in the Phase 3 COVE study.” The Pfizer/BioNTech information release makes less precise assertions about co-morbidities and other risk factors, to the effect that “Approximately 42% of global participants and 30% of U.S. participants have racially and ethnically diverse backgrounds, and 41% of global and 45% of U.S. participants are 56-85 years of age.” Other factors, such as geographic distribution of the trial subjects, may also explain the discrepancy.

From a comparison of the posteriors and credible regions for the two trials, it would appear that Moderna’s trial was better adapted to the purpose of assessing protective efficacy against severe COVID-19 than was the case with the Pfizer/BioNTech trial. The efficacy is *much* better-constrained in the Moderna trial than is the case for the Pfizer/BioNTech trial — Moderna’s 90% credible region is *e* ∈ [0.917, 1.0] in contrast to Pfizer/BioNTech’s 90% credible region of *e* ∈ [0.452, 0.993] Note that the fact that Moderna’s severe efficacy posterior peak is at *e* = 1.0 whereas Pfizer/BioNTech’s is not is merely due to the accident that *N*_*V*_ = 0 for the Moderna trial, as opposed to *N*_*V*_ = 1 for the Pfizer trial. As should be clear from the form of the reduced likelihood (Equation 2.5), the happenstance of observing 0 rather than 1 produces this posterior shape, which should not be overinterpreted. The most reasonable takeaway from this comparison is that the Pfizer severe COVID -19 efficacy is quite poorly constrained, but the overlap in the respective 90% credible regions again suggests that it is entirely possible on present data that the severe COVID-19 efficacies of the two vaccines are the same.

### 3.3. Prior Choice

To test robustness against prior choice, we select a prior that is somewhat informative compared to a uniform prior:

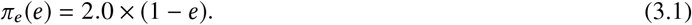

We refer to this prior as the “show-me” prior, because it embodies skepticism about values of *e* near 1.0, and insistence on very informative data before conceding that such values are plausible.

With the “show-me” prior, the results from the Bayesian analysis are changed as reflected in Table 3.

**Table 3:**
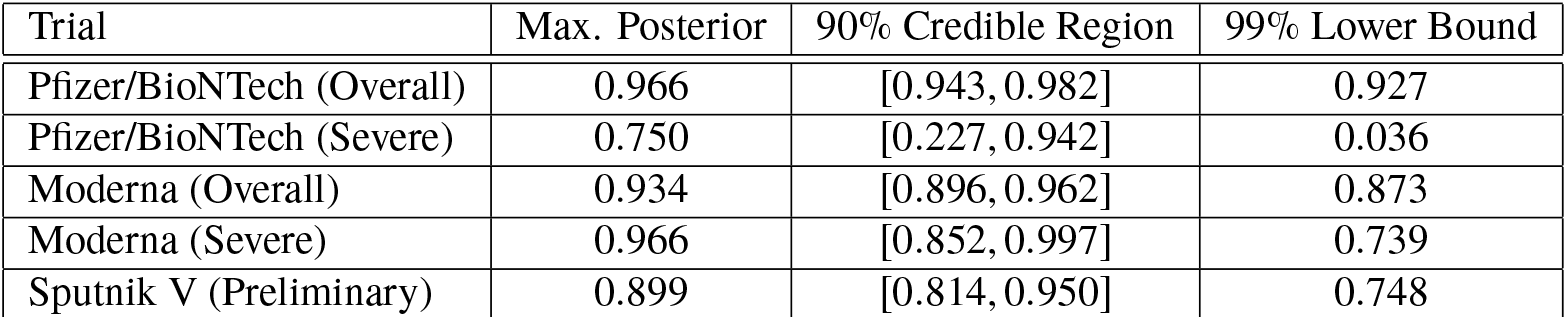
Summary of Bayesian efficacy estimation for SARS-CoV-2 vaccine trials assuming the “show-me” prior.

| Trial | Max. Posterior | 90% Credible Region | 99% Lower Bound |
| --- | --- | --- | --- |
| Pfizer/BioNTech (Overall) | 0.966 | [0.943, 0.982] | 0.927 |
| Pfizer/BioNTech (Severe) | 0.750 | [0.227, 0.942] | 0.036 |
| Moderna (Overall) | 0.934 | [0.896, 0.962] | 0.873 |
| Moderna (Severe) | 0.966 | [0.852, 0.997] | 0.739 |
| Sputnik V (Preliminary) | 0.899 | [0.814, 0.950] | 0.748 |

A comparison of the results in Tables 2 and 3 shows that the final results for the Pfizer/BioNTech and Moderna trials are barely affected by the change — the modifications are all in the third significant figure of the confidence regions and the posterior maxima. The modifications to the Sputnik V estimates are somewhat larger, but still modest — all in the second significant digit. Again, this is to be expected, in consequence of the lower numbers and broader likelihood for this preliminary trial data. The Moderna Severe results also vary modestly, in the second significant digit. Here the biggest change is that the “show-me” prior drives the posterior to 0 at *e* = 1.0, so the max-posterior point is moved to 0.966 instead. For these kinds of trial numbers, it is clear that even a rather extreme prior such as the “show-me” prior cannot affect the results of the analysis very substantially.

The largest change is in the Pfizer/BioNTech Severe results, where the changes are in the most significant digit. This is hardly surprising, since the likelihood shape (which is also the posterior shape in the top-right panel of Figure 3.1) is not even arguably well-concentrated in comparison to the “show-me” prior. For this kind of small-number data, more careful consideration of prior choice is clearly necessary.

## 4. Discussion

As illustrated in §3, the reduced likelihood approach leads to an easily-implemented 1-dimensional posterior density on the unit interval for the vaccine efficacy. Simple and intuitive results are easily extractable from such posteriors, and with the numbers characteristic of vaccine trials such as the three considered above, the overall efficacy estimates are quite robust to prior choice.

The most cogent take-away conclusion from the Bayesian analysis in §3 appears to be that on present data, the three vaccines for SARS-CoV-2 cannot be said to differ greatly in their efficacies. The differences are largely in the degree to which those efficacies are constrained by their respective trial data.

More thought needs to be given to prior choice in the event where subsamples have low counts. In the Pfizer/BioNTech Severe case, where there were 10 total infections, use of a skeptical prior such as the “show-me” prior can at least provide some conservatism to estimates. In such cases it is possible that more principled prior choice approaches, such as the reference prior approach advocated in [8] could lead to better-constrained, but still reasonable results. This is of course true irrespective of whether one adopts the reduced likelihood approach advocated here or another standard Bayesian nuisance parameterization.

Some generalization of the approach is possible: to test time-varying immunogenesis (whether short-term ramping up of immune response, or long-term decline of same, as in e.g. [16]), it is straightforward to introduce an empirical parametrized time dependence to the efficacy, i.e. *e* = *g*(*t* ; *θ*), where *θ* are parameters. If the data is in time bins, then the time-dependence *g* (*t* ; *θ*) could be integrated with respect to *t* over each time bin, and the likelihood computed as the product of the individual bin reduced likelihoods, which are now functions of *θ*. The resulting analysis no longer leads to simple manipulations of 1-dimensional posterior distributions on the unit interval, and MCMC methods may be advisable, depending on the dimensionality of *θ*. However, one benefit is that the nuisance parameter is still banished from the analysis, so that the parameter space has its dimensionality reduced by 1, and is limited to the empirical parameters of interest.

## Data Availability

The data utilized in this manuscript has been publically released by the companies involved.

https://www.pfizer.com/news/press-release/press-release-detail/pfizer-and-biontech-conclude-phase-3-study-covid-19-vaccine

https://investors.modernatx.com/news-releases/news-release-details/moderna-announces-primary-efficacy-analysis-phase-3-cove-study

https://sputnikvaccine.com/newsroom/pressreleases/second-interim-analysis-of-clinical-trial-data-showed-a-91-4-efficacy-for-the-sputnik-v-vaccine-on-d/

## Acknowledgements

This material is based upon work supported by the U.S. Department of Energy, Office of Science, under contract number DE-AC02-06CH11357. The author would like to acknowledge valuable clarifying conversations with William Fiske Parker (University of Chicago), Anne Sperling (University of Chicago), Aaron Esser-Kahn (University of Chicago), and Gordon D. Pusch (Argonne National Laboratory).

## Government License

The submitted manuscript has been created by UChicago Argonne, LLC, Operator of Argonne National Laboratory (“Argonne”). Argonne, a U.S. Department of Energy Office of Science laboratory, is operated under Contract No. DE-AC02-06CH11357. The U.S. Government retains for itself, and others acting on its behalf, a paid-up nonexclusive, irrevocable worldwide license in said article to reproduce, prepare derivative works, distribute copies to the public, and perform publicly and display publicly, by or on behalf of the Government. The Department of Energy will provide public access to these results of federally sponsored research in accordance with the DOE Public Access Plan. http://energy.gov/downloads/doe-public-access-plan.

## Footnotes

1 Given two posteriors *π*_1_(*e*_1_), *π*_2_(*e*_2_) over the efficacies *e*_1_ and *e*_2_ of two vaccines it would be a simple matter to compare the efficacies by computing a posterior density over the random variable Δ*e* ≡ *e*_2_ − *e*_1_, by implementing a discretized quadrature of the integral *π* (Δ*e*) = ∫ *de*_1_ *π*_1_(*e*_1_)×*π*_2_(*e*_1_+Δ*e*). We do not carry out such a comparison in this work.

## References

[1] M. E. Halloran, I. M. Longini, C. J. Struchiner, I. M. Longini, Design and analysis of vaccine studies, Vol. 18, Springer, 2010.

[2] S. E. Chick, D. C. Barth-Jones, J. S. Koopman, Bias reduction for risk ratio and vaccine effect estimators, Statistics in medicine 20 (11) (2001) 1609–1624.

[3] D. Katz, J. Baptista, S. Azen, M. Pike, Obtaining confidence intervals for the risk ratio in cohort studies, Biometrics (1978) 469–474.

[4] R. T. O’Neill, On sample sizes to estimate the protective efficacy of a vaccine, Statistics in Medicine 7 (12) (1988) 1279–1288.

[5] A. Agresti, B. A. Coull, Approximate is better than “exact” for interval estimation of binomial proportions, The American Statistician 52 (2) (1998) 119–126.

[6] S. E. Vollset, Confidence intervals for a binomial proportion, Statistics in medicine 12 (9) (1993) 809–824.

[7] H. Chu, M. E. Halloran, Bayesian estimation of vaccine efficacy, Clinical Trials 1 (3) (2004) 306–314.

[8] S. Laurent, C. Legrand, A bayesian framework for the ratio of two poisson rates in the context of vaccine efficacy trials, ESAIM: Probability and Statistics 16 (2012) 375–398.

[9] J. M. Bernardo, Reference posterior distributions for bayesian inference, Journal of the Royal Statistical Society: Series B (Methodological) 41 (2) (1979) 113–128.

[10] A. Gelman, J. B. Carlin, H. S. Stern, D. B. Dunson, A. Vehtari, D. B. Rubin, Bayesian data analysis, CRC press, 2013.

[11] Pfizer, Inc./BioNTech Inc., Pfizer and BioNTech Conclude Phase 3 Study of COVID-19 Vaccine Candidate, Meeting All Primary Efficacy Endpoints. URL https://www.pfizer.com/news/press-release/press-release-detail/pfizer-and-biontech-conclude-phase-3-study-covid-19-vaccine

[12] Moderna, Inc., Moderna Announces Primary Efficacy Analysis in Phase 3 COVE Study for Its COVID-19 Vaccine Candidate and Filing Today with U.S. FDA for Emergency Use Authorization. URL https://investors.modernatx.com/news-releases/news-release-details/moderna-announces-primary-efficacy-analysis-phase-3-cove-study

[13] R. D. I. Fund, Second interim analysis of clinical trial data. URL https://sputnikvaccine.com/newsroom/pressreleases/second-interim-analysis-of-clinical-trial-data-showed-a-91-4-efficacy-for-the-sputnik-v-vaccine-on-d/

[14] C. R. Harris, K. J. Millman, S. J. van der Walt, R. Gommers, P. Virtanen, D. Cournapeau, E. Wieser, J. Taylor, S. Berg, N. J. Smith, R. Kern, M. Picus, S. Hoyer, M. H. van Kerkwijk, M. Brett, A. Haldane, J.F. del R’ıo, M. Wiebe, P. Peterson, P. G’erard-Marchant, K. Sheppard, T. Reddy, W. Weckesser, H. Abbasi, C. Gohlke, T. E. Oliphant, Array programming with NumPy, Nature 585 (7825) (2020) 357–362. doi:10.1038/s41586-020-2649-2. URL https://doi.org/10.1038/s41586-020-2649-2

[15] J. D. Hunter, Matplotlib: A 2d graphics environment, Computing in Science Engineering 9 (3) (2007) 90–95.

[16] V. A. Morrison, G. R. Johnson, K. E. Schmader, M. J. Levin, J. H. Zhang, D. J. Looney, R. Betts, L. Gelb, J. C. Guatelli, R. Harbecke, et al., Long-term persistence of zoster vaccine efficacy, Clinical infectious diseases 60 (6) (2015) 900–909.

